# Genetic analysis and multimodal imaging identify novel mtDNA 12148T>C leading to multisystem dysfunction with tissue-specific heteroplasmy

**DOI:** 10.1101/2023.11.03.23297854

**Authors:** Kinsley Belle, Alexander Kreymerman, Nirmal Vadgama, Marco H. Ji, Sandeep Randhawa, Juan Caicedo, Megan Wong, Stephanie P. Muscat, Casey A. Gifford, Richard T. Lee, Jamal Nasir, Jill L. Young, Gregory Enns, Ioannis Karakikes, Mark Mercola, Edward H. Wood

## Abstract

Patients with mitochondrial disorders present with clinically diverse symptoms, largely driven by heterogeneous mutations in mitochondrial-encoded and nuclear-encoded mitochondrial genes. These mutations ultimately lead to complex biochemical disorders with a myriad of clinical manifestations, often accumulating during childhood on into adulthood, contributing to life-altering and sometimes fatal events. It is therefore important to diagnose and characterize the associated disorders for each mitochondrial mutation as early as possible since medical management might be able to improve the quality and longevity of life in mitochondrial disease patients. Here we identify a novel mitochondrial variant in a mitochondrial transfer RNA for histidine (mt-tRNA-his) [m.12148T>C], that is associated with the development of ocular, aural, neurological, renal, and muscular dysfunctions. We provide a detailed account of a family harboring this mutation, as well as the molecular underpinnings contributing to cellular and mitochondrial dysfunction. In conclusion, this investigation provides clinical, biochemical, and morphological evidence of the pathogenicity of m.12148T>C. We highlight the importance of multiple tissue testing and in vitro disease modeling in diagnosing mitochondrial disease.

## INTRODUCTION

Mitochondrial diseases affect 1:5,000 people^1^, however with 1:200 people carrying a known deleterious mitochondrial DNA (mtDNA) mutation^2^, evidence suggests that mitochondrial mutations may have an even greater frequency of disease burden. Many of these pathogenic mtDNA mutations have been identified in mitochondrial transfer RNAs (mt-tRNAs), with an overwhelming 60% of mitochondrial disorders associated with mutations in mt-tRNAs in contrast to mutations affecting other components of mitochondria^3^. The higher frequency of mt-tRNA mutations is thought to potentially be due to a stem-loop structure formed during mtDNA replication, which could then expose a single stranded portion of mt-tRNA genes to a greater rate of oxidative damage compared to other mtDNA regions^4,5^. A greater rate of mt-tRNA mutation-linked mitochondrial disease may also be largely driven by the fact that a single mutated mt-tRNA can potentially alter the translation of all the mtDNA encoded proteins, leading to a greater potential for metabolic disruptions by influencing events across the entire electron transport chain (ETC). As result, detectable mt-tRNA mutations are marked as “possible pathogenic” in sequencing studies, with 458 mt-tRNA mutations identified to date. Yet, elevating the status of a mt-tRNA mutation from a “possible” to a “confirmed” pathogen remains challenging in a clinical setting, with currently only about 98 pathogenic mutations “confirmed”-with likely many others yet to be discovered or defined as pathogenic^6^.

The challenges in detecting and diagnosing a mtDNA mutation as disease causing is in part because of mitochondrial heteroplasmy, a scenario in which mutations in mtDNA can manifest at variable amounts per cell and/or tissue^7^. This also means that mtDNA mutations in a heteroplasmic state have the potential to be missed during clinical diagnosis, as tissues carrying mtDNA mutations and experiencing symptoms may not be readily accessible for biopsy, e.g., muscular, cardiac, nervous, and renal systems. In fact, clinical assessments of heteroplasmy are often performed on blood and buccal swabs, although these tissues do not accurately reflect the heteroplasmy in affected organs^7,8^. Furthermore, the most impacted tissues in these diseases, neural and cardiac tissues, are typically unavailable for *in vitro* functional studies. Thus, to overcome this obstacle, induced pluripotent stem cell (iPSC) models have emerged as a possible tool to model tissues and the functional impact of a particular mtDNA mutation and its heteroplasmic state on these tissues.

Herein, we describe a family harboring a unique mutation of the mt-transfer RNA for histidine (mt-tRNA-his) [m.12148T>C] (Eurasian population frequency 0.002378%, total population frequency 0.001635%) with a mitochondrial syndrome affecting multiple organ systems, previously reported but not substantiated as disease-contributing^9,10^. Through in-depth characterization of this novel mt-tRNA syndrome, we highlight the importance of multiple tissue site sampling and sequencing with single cell analysis for identifying mutation carriers. We also used fibroblasts and iPSC-differentiated cardiac and neuronal cells to show the consequences of this mt-tRNA mutation on cell function. Overall, this study provides the first characterization of the pursuant disease in carriers of m.12148T>C and verifies this mutation as the probable cause of this patient cohort’s disease, changing this variant’s status from “possibly pathogenic^11^” to “confirmed pathogenic” and setting a precedent for monitoring patients for potential mitochondrial disease if the m.12148T>C mutation is detected at any level.

## MATERIALS AND METHODS

### Patient Data

This study was approved by the institutional review board of Stanford University. All subjects gave written informed consent before undergoing evaluation, testing, and sequencing.

### Cell Line Establishment

To establish fibroblast lines, fresh ¾ inch skin punch biopsies were first quickly dipped in 70% ethanol, and then cut into smaller pieces prior to seeding onto gelatin-coated 6 well tissue culture plates with a cover slip placed on top to aid in attachment. Skin punches were then cultured in DMEM, supplemented with 10% FBS and antibiotics-antimycotics (Gibco, 15240096) for the first five days, then without antibiotics. Significant fibroblast growth was observable at the edges of punch biopsies by day 14, at which point cells were trypsinized and seeded into new gelatin-coated 6 well plates for subsequent experimental expansion, sequencing investigations, and/or cryopreservation in Bambanker (Lymphotec Inc.). All fibroblasts were maintained at below 6 passages.

Uroepithelial cells were isolated and grown from fresh urine. To accomplish this, urine was centrifuged at 500xg for 5 minutes. The pellet was then washed twice with PBS, by resuspending in PBS and centrifuging twice more. The resulting pellet of cells was then resuspended in Renal Epithelial Growth Medium (Lonza, CC-3190) and allowed to grow as previously described^12^. The cells then underwent qPCR heteroplasmic analysis and reprogramming.

Peripheral mononucleated blood cells (PBMCs) were separated from whole blood as described in the protocol from the Lymphoprep/Sepmate system (StemCell technologies, 85450 and 07801). Briefly, whole blood was diluted in PBS + 2% FBS solution, layered onto the Lymphoprep gradient in Sepmate tubes, and spun at 1200xG for ten minutes. The separated PBMCs were then poured into new tubes, washed twice in a PBS + 2% FBS solution, and pelleted at 300g for 8 min. Samples were then either grown out for subsequent experiments, cryopreserved, or used in sequencing studies.

### Single Cell Sort for Outgrowth and Heteroplasmy Determination

To initiate clonal expansion of our lines and to determine single cell heteroplasmy, cells were sorted using BD biosciences FACS ARIA III based on scatter and then seeded at one cell per well into a 96 well PCR or cell culture plate. To determine heteroplasmy at the single cell level, cells were sorted into PCR plates and heated to 100°C, mitochondrial genomes were amplified using REPLI-g Mitochondrial DNA Kit (Qiagen, 151023), and the resulting material was tested for heteroplasmy by qPCR. Cells plated into 96 well culture plates were expanded from a single cell level until confluency and then passaged into 12 or 24 well plates for further expansion, experimental use, or in heteroplasmy qPCR assessment, as in single cell experiments.

### Induced Pluripotent Stem Cell (iPSC) Generation

iPSC lines derived from patient primary cells were created as per standard protocol using CytoTune™-iPS 2.0 Sendai Reprogramming Kit (A1657). Cells were seeded, transduced with SENDAI virus, and grown with mTESR e8. On day 14 post transduction, single colonies were clonally selected and grown. After 5-7 days cells were reseeded on feeder-free substrates and propagated to passage 20 for all experiments.

### Differentiation of Dopaminergic neurons

Dopaminergic neuronal cultures were initiated with the formation of neural aggregates from the described isogenic iPSC lines. Dissociated single cells were loaded into AggreWell™800 to form neural aggregates. At this point, cells were treated with neural induction media. Dual-SMAD inhibition was initiated with Noggin (Peprotech 120-10C)/SB431542 and purmorphamine (Peprotech 4831086) was added to specify midbrain lineage. At day 5 aggregates were plated on to laminin substrate and expanded with bFGF (Peprotech 100-18B), Forksolin, and SHH/FGF8 (Peprotech 100-25A) to further specify midbrain lineage. Three weeks after the start of this protocol, cultures were re-plated onto PDL/LAMININ in neurobasal N2/B27 media with FGF8, SHH, TGFβ-3 (Peprotech 100-36E), BDNF (Peprotech 450-02), GDNF (Peprotech 450-10), Forksolin, Ascorbic Acid. FGF8, and bFGF were removed from culture on day 27, and SHH was reduced. Cells were fed with half media changes every other day and were able to be cultured for long periods of time by replating on fresh PDL/Laminin.

### Differentiation of Cardiomyocytes

Cardiomyocytes were derived using the metabolic maturation protocol previously described^13^. Confluent lines were cultured in RPMI (Gibco 11875101) supplemented with B27 (without insulin) plus 6 μΜ CHIR. CHIR concentrations were reduced over the next 2 days by the additional media. On day 3, the media was changed and supplemented with 2uM Wnt-C59. On day 7, the media was then changed to RPMI supplemented with B27 now with insulin added. Thereafter, the media was changed every other day until day 11. Cells were then placed in a starvation state: RPMI without B27 or glucose for 3 days. On day 15, cells were replated in RPMI + B27+insulin. On day 17, a final starvation state was induced for three days. On day 20, cells were cultured in RPMI+B27+insulin, and then cultured in maturation media from that point on (as described in Feyen et al).

### Cardiomyocytes Contractility Measurements

HiPSC-CMs were plated on Geltrex at 40,000 cells per well of a 384-well plate (Greiner BioOne) in 75µl of RPMI/B27+insulin containing 10% KOSR and ROCK inhibitor Y-27632 (Tocris, 1253). The next day media was changed to RPMI/B27+insulin final volume 90µl and cells were maintained for a minimum of 7 days prior to imaging. For the analysis, 45µl of media was removed and hiPSC-CMs were loaded with 45µl 2x TMRM (T668, ThermoFisher Scientific) with Hoechst 33258 (ThermoFisher Scientific). After 10min incubation in a 37°C 5% CO2 incubator, cells were washed by removing 90μL of media and replacing it with 90μL RPMI/B27+insulin. After a 2hr recovery period, timed series images were acquired automatically using the IC200 KIC instrument (Vala Sciences, California, USA) at an acquisition frequency of 50Hz for a duration of 10sec. The contractility image analysis, using TMRM signal derived vectorized points of movement through a time series and physiological parameter calculations were conducted, as previously described^14^. D-peak represents peak divergence or the average peak of contraction across a 10sec recording, used to graphically represent contractile changes relative to controls^15^.

### qPCR Determination of Heteroplasmy

To determine heteroplasmy, amplification-refractory mutation system (ARMS) qPCR primers were designed to selectively amplify mutated mtDNA. Two pairs of sequence specific primers were designed with penultimate mismatches, and base changes to decrease possible hairpin formation and primer dimerization^16^. Multiple primer sets were assayed with the following being selected for use: COMMON FWD 5’-AACACAATGGGGCTCACT-3’, WT REV 5’-GACTCACGATCTGATGTTTTGGTTATA-3’, and m12148C REV5’-GACTCACGATCTGATGTTTTGGTTATG-3’. Samples to be assayed were prepared in replicate pairs, and each qPCR assay was done in triplicate with required controls, using BIORAD iTaq Universal SYBR Green Supermix (Biorad, 1725120). Cycle threshold (Ct) values were then used to calculate the percent heteroplasmy using a previously established method, 1/[1+(1/2)^(CtWT-CtMut)]*100^17^.

### Whole-exome sequencing

Whole-exome sequencing was performed using the Illumina PE150 (Illumina Inc., USA) with average insert lengths of 350 bp. Sequence alignment to the human reference genome (UCSC hg19), followed by variant calling and annotation. In brief, this involved alignment with Burrows-Wheeler Aligner (BWA), removal of PCR-duplicates with Picard Tools, followed by sample-paired local realignment around indels and germline variants identified by HaplotypeCaller, according to the Genome Analysis Toolkit (GATK) best practices.

### Variant filtering and selection

The variants were categorized according to gene annotation, population allele frequency, functional prediction, and clinical interpretation. The raw list of SNVs and indels was annotated using ANNOVAR^16^.

Variants were classified based on their mutational characteristics, position in the genome, and allele frequency. *In silico* prediction of pathogenicity was performed using CADD^18^ and REVEL^19^ and conservation of nucleotides was scored using GERP++^17^. Population allele frequencies were obtained from 1000Genomes, and gnomAD^20^. Variants –either heterozygous or homozygous-that were shared between the affected individuals were retained for downstream analysis.

Variants in splicing regions, 5′-UTR, 3′-UTR, and protein-coding regions, such as missense, frameshift, stop loss, and stop gain mutations, were considered. Variants were retained if they had a minor allele frequency (MAF) < 0.001. Rare variants were ranked according to a REVEL score above the default threshold of 0.5, a CADD score greater than 20, and GERP++ score greater than 2. Variants with clinical significance as benign or likely benign according to the ClinVar dataset were removed.

### Network analysis

A functional enrichment analysis of the prioritized candidate genes was performed using VarElect^21^. This tool uses the deep LifeMap Knowledgebase to infer the “direct” or “indirect” association of biological function between variant containing genes and the queried phenotypes— e.g., retinal dystrophy, myopathy, pseudohypoparathyroidism, hypertension, focal segmental glomerulosclerosis, movement disorder, intracranial calcifications, and sensorineural hearing loss. A direct association is determined if studies indicate that the gene in question directly affects disease development. An indirect association is based on factors such as shared pathways, protein–protein interaction networks, and multiple publications. VarElect generates a score that can be used as an indication of the strength of the connection between a variant/gene and the queried phenotypes. The score helps to rank and prioritize the list of queried genes by relevance to the disease.

### Image Analysis of Mitochondrial Heteroplasmy in Fibroblasts

Images of wild-type fibroblasts and heteroplasmic mtDNA 12148T>C fibroblasts were captured with confocal microscopy and analyzed. The high-resolution images are 1,900 x 1,900 pixels and display single cells were fixed in a 4% PFA PBS solution for 15min, permeabilized, and stained with DAPI to reveal nuclei. To reveal mitochondrial networks and cell membranes, cells were exposed to baculoviruses expressing fluorescent proteins (CellLight C10600 and C10608, Thermofisher Scientific), which label the mitochondrial and cell membranes respectively, prior to fixation and permeabilization. We followed an image-based profiling methodology to analyze the sets of images^22^. We then quantified the mitochondrial structures of all single cells by cropping overlapping regions of 300 x 300 pixels in areas of the images where the mitochondria had highest activation. This resulted in approximately 32 overlapping regions where the mitochondrial structures were sampled for quantification. A total of 418 mitochondrial regions were then extracted from the images. Morphological features of each of the regions were computed using a pre-trained convolutional neural network^22,23^ resulting in a 1,536-dimensional vector representing the structural content of the image. Before feature computation, the mitochondrial channel was normalized using Contrast Limited Adaptive Histogram Equalization (CLAHE). The software used to compute deep learning features was DeepProfiler. The feature extraction procedure resulted in a matrix of 418 regions with 1,536 features each, representing the regions extracted from the images.

### Biochemical analysis of mitochondrial enzymatic activity

Complex I and Complex IV activity were measured using Complex I Enzyme Activity Microplate Assay Kit (ab10972, Abcam) and Complex IV Human Specific Activity Microplate Assay kit (ab109910, Abcam). For Complex I measurements, fibroblasts were homogenized, protein was detergent-extracted, and quantified using standard BCA methods (23225, Pierce BCA Protein Assay Kit). Extracts of lines and appropriate controls were then incubated for 3 hours in appropriate buffers. Assay solution was added and OD at 450nm was measured every 30 seconds for 30 minutes. Rate and slope were then calculated for all samples using: Rate mOD/min = Abs 1 – Abs 2 (mOD @ 450nm) Time 1 – Time 2 (min).

For Complex IV measurements, fibroblasts were homogenized, protein was detergent-extracted, and quantified using standard BCA methods. Samples and appropriate controls were loaded onto the plate and incubated for 3 hours. Assay solutions were then added and OD at 550nm was measured at 1-minute intervals for 2 hours to determine enzyme activity: Rate mOD/min = Abs 1 – Abs 2 (mOD @ 550nm) Time 1 – Time 2 (min). Then, to determine the total amount of bound complex protein in the plate, antibodies to Complex IV were applied to the same wells used in the enzyme activity assay. Bound protein was reacted according to kit parameters and detected via OD measures at 405nm every minute for 30 minutes. Bound complex concentration relative to total protein loaded was calculated within the linear portion of the curve and used to normalize Complex IV enzyme activity.

### Analysis of mitochondrial respiration

Mitochondrial respiration was assayed using the Seahorse XFe96 Extracellular Flux Analyzer (Agilent Technologies). Derived cell lines were plated into XF96 cell culture microplates and allowed to grow for 48hrs. Prior to the assay, cells were equilibrated in XF DMEM assay media supplemented with 25mM glucose, 2mM Glutamax, 1mM pyruvate (103680-100, Agilent Technologies). Respiration parameters were then assayed by using the Mito Stress Test Kit (103708-100, Agilent Technologies) and by applying oligomycin (1.5 µM), carbonyl cyanide 4-(trifluoromethoxy) phenylhydrazone (FCCP, 2 µM) and rotenone/antimycin A (0.5 µM). Basal respiration rates, maximal respiration (respiration in response to FCCP) and reserve capacity (maximal respiration – basal respiration) were then established.

## RESULTS

### Clinical Descriptions and Genetic Workup

**Patient 1**, the proband, was a former university graduate student who succumbed to her illness in her 30s. She had been healthy in early childhood, with normal vision and hearing evaluated at ages 5-10. However, by the age of 6-10 she developed progressive sensorineural hearing loss requiring cochlear implants in 2004 at ages 10-15. Imaging prior to the cochlear implant surgery revealed areas of intracranial calcifications (**Figure 1A**). A muscle biopsy (**Figure 1B**) as part of her diagnostic evaluation revealed abnormal mitochondria, which were also visualized under electron microscopy (**Figure 1C**). A kidney biopsy obtained due to clinical symptoms of renal disease in early adulthood revealed focal segmental glomerular sclerosis (FSGS) along with mitochondrial abnormalities (**Figure 1D and E**). Additional issues that developed over the last decade of her life included: muscle wasting and weakness, chronic renal failure, hypertension, retinal dystrophy (**Figure 1F**), petit mal seizures, ataxia, parkinsonian movement disorder, cognitive decline, and autonomic dysfunction. She experienced a stroke-like episode with transient aphasia and confusion that resolved, but thereafter she rapidly declined and died ten days later.

**Figure 1.**
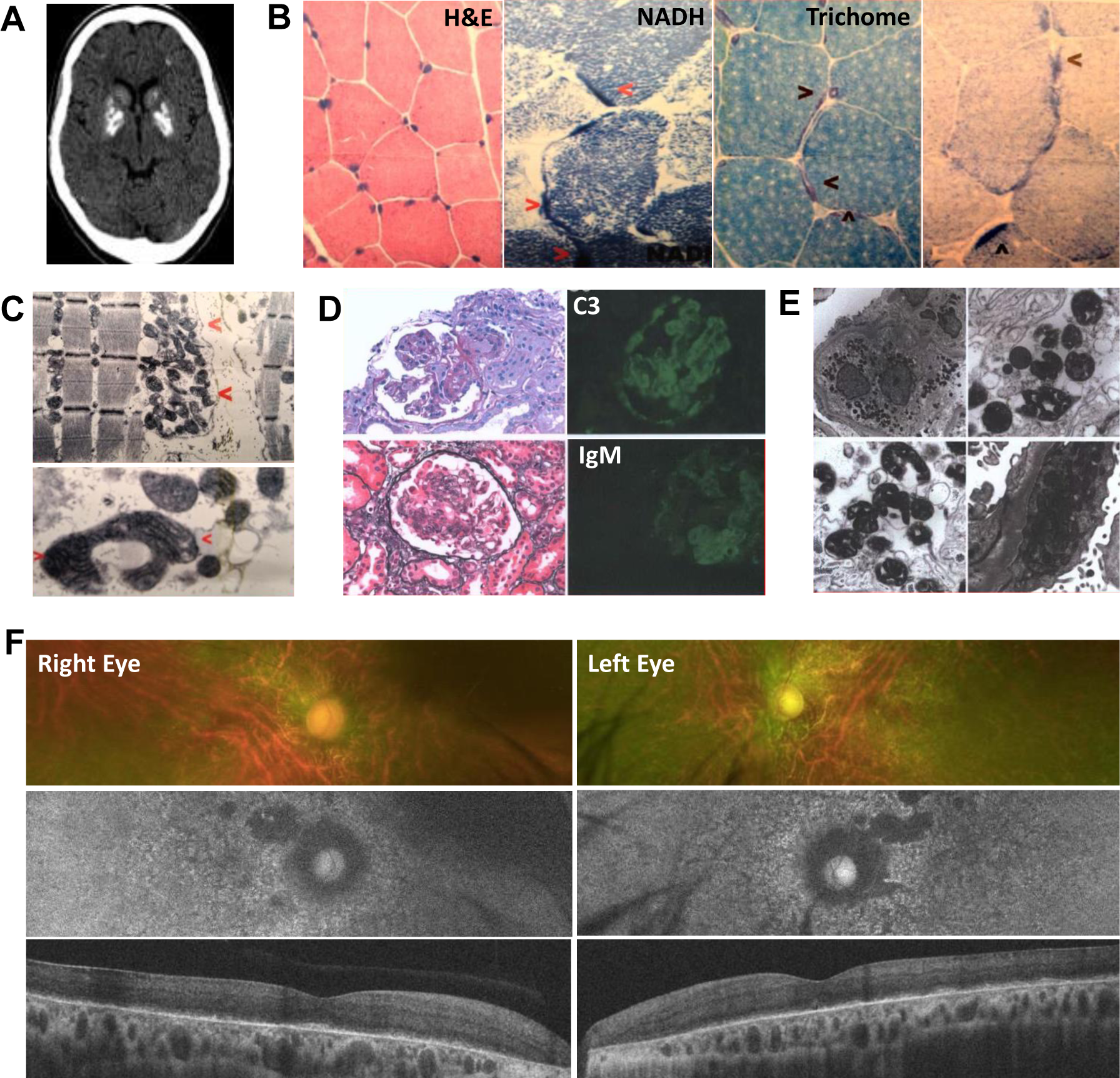
Multiple tissue dysfunction assessed in the proband, highlights medical evidence of mitochondrial disease. **(A)** At ages 11-16 the proband received a CT scan revealing significant basal ganglia and cerebellar calcifications, prompting further testing. **(B)** Muscle biopsy specimens of proband acquired at age 11-16 showing haematoxylin and eosin (H&E) staining of muscle, along with increased nicotinamide adenine dinucleotide dehydrogenase (NADH) staining (red arrows), increase trichome staining (blue arrows), and increased succinic dehydrogenase (SDH) staining, all indicative of mitochondrial disease. **(C)** Samples acquired the same year were examined by transmission electron microscopy (TEM), display abnormal mitochondrial morphology (red arrows) in muscle with two representative sections (zoomed out-top and zoomed in-bottom panel). (**D)** At age 11-16 samples from a renal biopsy were acquired. H&E staining presenting collapsed glomeruli, with antibody staining showing increased complement 3 (C3) deposits and Immunoglobulin M (IgM) staining revealing inflammation. **(E)** TEM imaging of renal tissue, revealing dysmorphic features and abnormal mitochondria morphology. **(F)** Multimodal retinal imaging in both eyes reveals symmetrical waxy disc pallor (top panel), significant loss of retinal pigment epithelial (RPE) cells on fundus autofluoresence (FAF) (middle panel), and outer retinal atrophy with loss of the ellipsoid zone (EZ) and interdigitation zone (IZ) on optical coherence tomography (OCT) (bottom panel).

Despite the abnormal mitochondrial morphology found in the 2004 muscle biopsy, the exome sequencing and metabolic testing available at that time failed to identify a specific defect. However, as newer technologies became available, targeted mtDNA sequencing performed in 2014 revealed a novel, presumed pathogenic, m.12148T>C heteroplasmic variant in the mitochondrial tRNA for histidine. This novel m.12148T>C variant was found to be present at a heteroplasmic level of 94% in skeletal muscle, 68% in urine sediment, and 35% in blood. In addition, mtDNA sequencing revealed that she was also carrying a known homoplasmic but benign mtDNA polymorphism (m.12937A>G, p.M201V in ND5), and a known homoplasmic variant m.961T>C (12s rRNA) associated with hearing loss in patients prescribed aminoglycosides without other systemic manifestations. This polymorphism was also found in grandparent, parent and sibling of proband. The grandparent and sibling have no clinical signs of deafness (**Table 1).**

**Table 1:** Cohort Clinical and mtDNA Observation.

|  | <u>Clinical Findings</u> | <u>MtDNA Variants</u> |
| --- | --- | --- |
| **Proband** | **Age:** 26-30<br>**Symptoms:** Cataracts, retinal dystrophy, hearing loss, myopathy, cognitive decline, parkinsonian movement disorder, ataxia, renal disease, hypertension, autonomic dysfunction | 1. m.12148T>C (tRNA-His) [Heteroplasmic]<br>2. m.961T>C (12s rRNA) [Homoplasmic]<br>3. m.12937A>G (p.M201V in ND5) [Homoplasmic, benign polymorphism] |
| **Parent of Proband** | **Age:** 56-60<br>**Symptoms:** Cataracts, hearing loss, cognitive decline | 1. m.12148T>C (tRNA-His) [Heteroplasmic]<br>2. m.961T>C (12s rRNA) [Homoplasmic] |
| **Sibling of Proband** | **Age:** 26-30<br>**Symptoms:** None | 1. Negative<br>2. m.961T>C (12s rRNA) [Homoplasmic] |
| **Grandparent of proband** | **Age:** 81-85<br>**Symptoms:** None | 1. Negative<br>2. m.961T>C (12s rRNA) [Homoplasmic] |

**Patient 2** is the parent of the proband, currently age 56-60 (**Figure 2A**). At age 1-5, she contracted bacterial meningitis (Haemophilus influenzae type b) and was treated with chloramphenicol, but otherwise had a generally healthy childhood and early adulthood, giving birth to the proband and an additional unaffected daughter. She initially had mtDNA sequencing performed from blood in 2008 in her early 40s, at which time she had mild hearing loss, and was found to be negative for mtDNA mutations. By her early 50s, she had developed severe bilateral sensorineural hearing loss, cataracts, and mild cognitive impairment, prompting her to repeat whole mtDNA sequencing at Baylor in 2017 which disclosed the m.12148T>C variant present in 10% heteroplasmy in urine. In 2020, we re-performed dedicated whole mtDNA sequencing which detected the m.12148T>C variant at 1% heteroplasmy in fibroblasts and 5% heteroplasmy in blood. We also confirmed the presence of the homoplasmic variant m.961T>C (12s rRNA), shown to be homoplasmic through the maternal line in both the affected and unaffected family members. (**Table 1**)

**Figure 2:**
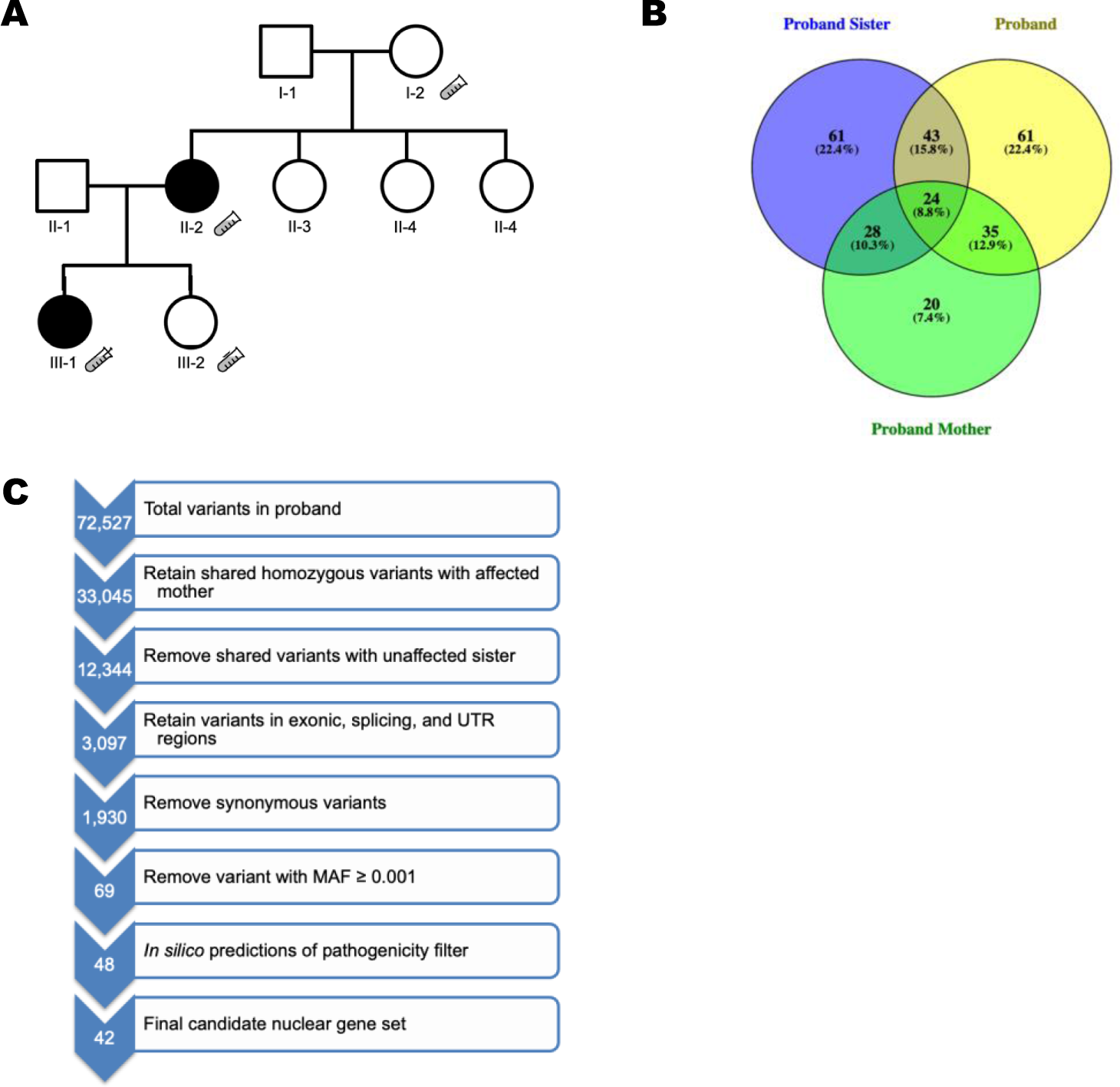
Genetic Analysis of Cohort highlights novel mtDNA tRNA-his M.12148T>C mutation. **(A)** Pedigree Cohort tube denotes Whole Exome Sequencing WES **(B)** Venn Diagram denotes shared and unshared variants between affected patients 35. **(C)** Variant Filtering yields 35 possible nuclear variants, Whole Exome sequencing yielded 11503 possible variants, filtering out known common variants reduced this to 187, subsequent phenotypic filtering yield 156 possible variants. Lastly familial filtering highlighted potential variants of interest, these variants were subsequently deemed not related to patient phenotype.

**Patient 3** is the 21-26-year-old unaffected younger sister of the proband. She had mtDNA sequencing performed from blood in 2008 and was found to be negative. She later had whole mtDNA sequencing re-performed in 2017, which was also negative. In 2019, we re-performed dedicated PCR-based mtDNA sequencing for the m.12148T>C variant and found that it was not detectable. As expected, we also confirmed the presence of the homoplasmic variant m.961T>C (12s rRNA) (**Table 1**).

**Patient 4** is the unaffected maternal grandparent of the proband, at age 80-85 is healthy and without symptoms suggestive of mitochondrial disease. She was tested for the m.12148T>C mutation from a buccal swab, which was undetectable at levels even below 1% heteroplasmy. In 2019, we re-performed dedicated PCR-based mtDNA sequencing for the m.12148T>C variant and found no detectable level of this variant. As with all other patients in this cohort, we also detected the known homoplasmic variant m.961T>C (12s rRNA) (**Table 1**).

Finally, to rule out other confounding nuclear mutations as disease sources, we performed whole-exome sequencing (WES) of the proband (affected), parent (affected) and sibling (unaffected). Pedigree mapping did not suggest an autosomal recessive model of inheritance. (**Figure 2A**). Thus, variant filtering was applied for a dominant mode of inheritance. (See methods). Finally, a variant was considered a disease-causing candidate if it was detected in both affected individuals, and absent in the unaffected. This filtering criteria yielded 42 putative genes with variants (**Fig. 2B-C**). We analyzed the 42 genes from the WES analysis, including *MT-TH* from the mitochondrial analysis (n = 43), using VarElect to correlate their functions with different aspects of the clinical phenotype. Results suggest that 18 targets were directly related to the phenotype, whereas 24 were indirectly related (**Table 2**). Among the unified results, *MT-TH* exhibited the highest phenotype association based on a VarElect score of 41.91 (**Table 2**), while the prioritized nuclear genes scored relatively low suggesting that they were not the leading cause of the proband’s symptoms. Thus, taken together, these findings suggest that the pathology identified in our patient cohort is likely due to the mitochondrial variant m.12148T>C, common to the proband and the proband’s parent.

**Table 2.** Top 10 candidate genes directly or indirectly associated with proband phenotypes ranked by VarElect. The score is an indication of the strength of the connection between the gene and the disease.

| Rank | Gene | Type | Matched Phenotypes | Direct / Indirect | Score |
| --- | --- | --- | --- | --- | --- |
| 1 | MT-TH | ncRNA | myopathy; hypertension; focal segmental glomerulosclerosis; intracranial calcifications; sensorineural hearing loss | Direct | 41.91 |
| 2 | RNASEH2C | Protein | hypertension; movement disease; intracranial calcifications; sensorineural hearing loss | Direct | 9.25 |
| 3 | KIRREL3 | Protein | hypertension; focal segmental glomerulosclerosis; sensorineural hearing loss | Direct | 7.97 |
| 4 | SCNN1A | Protein | hypertension | Direct | 5.98 |
| 5 | RAB11B | Protein | myopathy; pseudohypoparathyroidism; hypertension; intracranial calcifications; sensorineural hearing loss; retinal dystrophy; focal segmental glomerulosclerosis | Indirect | 3.71 |
| 6 | SIPA1 | Protein | retinal dystrophy; myopathy; hypertension; focal segmental glomerulosclerosis; sensorineural hearing loss; intracranial calcifications; pseudohypoparathyroidism | Indirect | 3.44 |
| 7 | SRGAP2 | Protein | retinal dystrophy; myopathy; hypertension; focal segmental glomerulosclerosis; sensorineural hearing loss; pseudohypoparathyroidism; intracranial calcifications | Indirect | 3.29 |
| 8 | ATP1A1 | Protein | hypertension | Direct | 3.01 |
| 9 | PDE4DIP | Protein | retinal dystrophy; myopathy; hypertension; sensorineural hearing loss; pseudohypoparathyroidism; focal segmental glomerulosclerosis | Indirect | 2.83 |
| 10 | ARHGEF26 | Protein | hypertension; focal segmental glomerulosclerosis; sensorineural hearing loss; retinal dystrophy; myopathy; pseudohypoparathyroidism; intracranial calcifications | Indirect | 2.36 |

### Bulk tissue and Single Cell Analysis of Mitochondrial DNA Heteroplasmy

With mitochondria sequencing confirming the presence of m.12148T>C as a potential source of disease, we then focused on identifying the heteroplasmic state of m.12148T>C in samples acquired from our disease cohort. Heteroplasmy has been shown to vary across different tissues, so we measured heteroplasmy in the proband across multiple tissues including blood, dermal fibroblasts, and urine sedimentary cells (the tissues sampled were obtained 5 years premortem). Heteroplasmy was determined utilizing allele refractory mutation system (ARMS)-based quantitative PCR and reflected as a percentage of amplified m.12148T>C copies relative to wild type mitochondrial DNA. This analysis revealed that all samples were positive for the m.12148T>C mutation but at varying levels of heteroplasmy, with fibroblasts reflecting ∼56%, blood 40%, and renal epithelial cells 68% heteroplasmy levels **(Table 3).** These values were similar to those observed in clinical assessments and as confirmed by mtDNA next-generation sequencing.

**Table 3.**
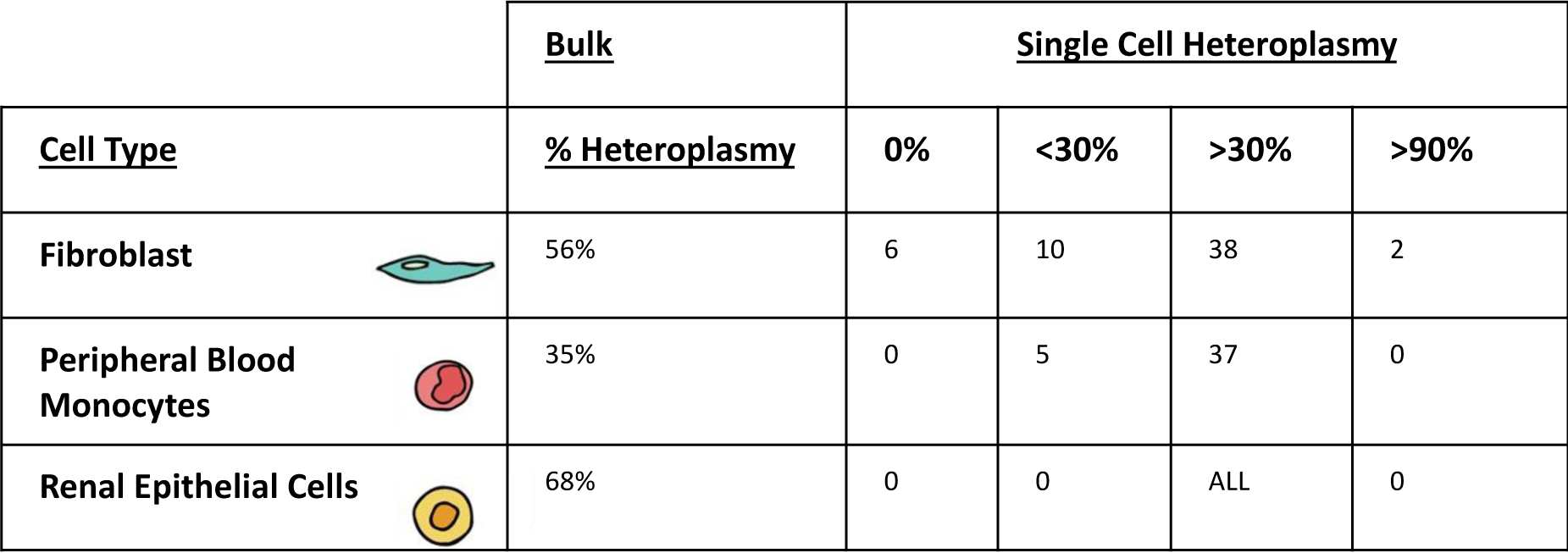
HeteroplasmyAcrossPatient Tissues.

| <u>Cell Type</u> |  | <u>Bulk</u> | <u>Single Cell Heteroplasmy</u> |  |  |  |
| --- | --- | --- | --- | --- | --- | --- |
|  |  | <u>% Heteroplasmy</u> | 0% | <30% | >30% | >90% |
| **Fibroblast** |  | 56% | 6 | 10 | 38 | 2 |
| **Peripheral Blood Monocytes** |  | 35% | 0 | 5 | 37 | 0 |
| **Renal Epithelial Cells** |  | 68% | 0 | 0 | ALL | 0 |

To gain insight into how heteroplasmy was distributed in the tissues sampled, we then followed the results from bulk sequencing and PCR with single cell analysis of heteroplasmy. This was accomplished through disassociation of cells carrying the m.12148T>C mutation, sorting one cell/well in a 96-well plate, and then assaying for m.12148T>C mtDNA load via ARMS-based quantitative PCR in single cells. This approach revealed that fibroblasts had a broad range of heteroplasmy with about 11% of cells carrying undetectable levels of heteroplasmy, 18% expressing <30% heteroplasmy, 68% expressing >30% heteroplasmy, and about 3% approaching homoplasmy (>99% heteroplasmy status). While in contrast, urine and blood cells exhibited a smaller range of heteroplasmy, with blood derived PBMCs showing about 12% of cells with <30% and about 88% of cells with >30% heteroplasmy, and urine captured cells mainly expressing heteroplasmy in the >30% range. The broad range of heteroplasmy observed in the fibroblast lines made it a useful tool for subsequent investigations. From the fibroblast lines we then derived isogenic wild type (WT) and heteroplasmic lines (HP) to measure the direct effect of m.12148T>C.

### Image Analysis of Mitochondrial Heteroplasmy in Fibroblasts

Mitochondrial health and function have been directly correlated to morphology, therefore we sought to explore how heteroplasmy of m12148T>C would affect morphology. Since the fibroblast lines had the greatest variance of heteroplasmy, we utilized the resultant lines to assess how heteroplasmy affects mitochondrial morphology, comparing high heteroplasmic (HP) expressing lines to lines with no detectable mutation or “Wild Type” (WT) lines. Observations of fluorescent labeled mitochondria in HP and WT lines highlighted differences in mitochondrial networks (Figure 3A). We then partitioned cell images into mitochondrial regions (red boxes) to increase experimental n from n=13 to n=418, to achieve increased significance (**Figure 3B)**. Next, we applied machine learning to identify the differences between the two cell lines. Principal Components Analysis (PCA) was utilized to reduce the dimensionality of features from 1,536 to 100 dimensions (**Fig. 3C**). In this reduced feature space, we performed multiple-hypothesis corrected Student t-tests to identify which of these dimensions are relevant to distinguish between HP and WT fibroblasts. When plotting the significantly different dimensions, we observed a consistent separation of mitochondrial patterns between the groups (**Figure 3C**), suggesting that mitochondrial morphology is affected by the presence of the m.12148T>C mutation. Finally, we tested whether the distribution of correlations between mitochondrial regions organized in three different groups: self-correlation of wild-types, self-correlation of heteroplasmic regions, and cross-correlation between wild-types and heteroplasmic regions. It was anticipated that the self-correlations would have high values and cross-correlations would have low values, meaning that there are consistent intra-group similarities and that there are significant inter-group differences. To test this hypothesis, we built the distribution of correlations between the three groups and used a Kruskal-Wallis nonparametric test assuming the null hypothesis that there is no difference between the distributions. The results are shown in (**Figure 3D)**, indicating that the differences observed between WT and HP mitochondrial regions are statistically significant (p-value < 0.0001).

**Figure 3:**
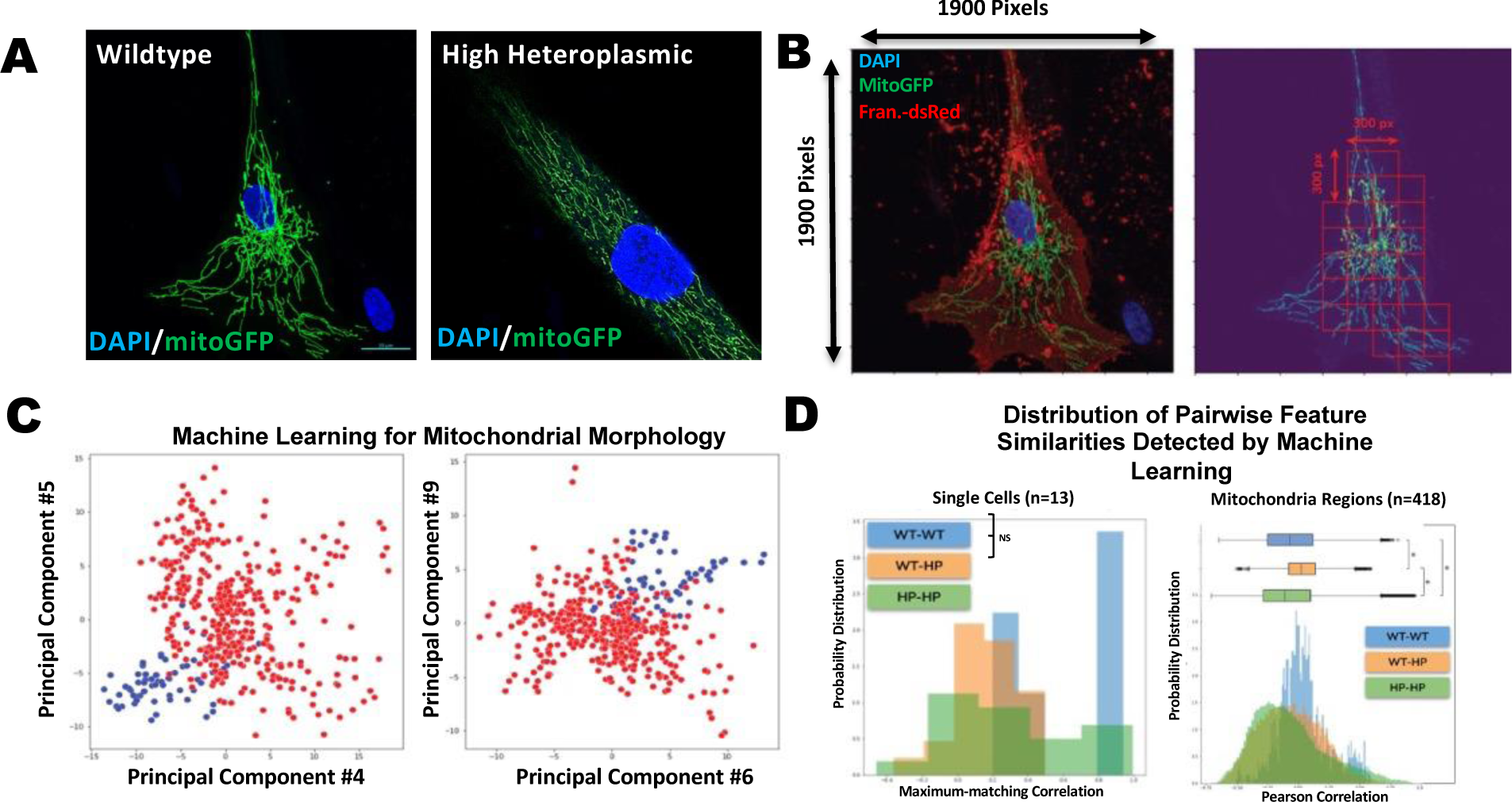
Mitochondrial Imaging. **(A)** High-resolution confocal images of fibroblasts included in this analysis. **(B)** Regions of the mitochondria channel extracted and analyzed in this study. The original resolution of confocal images is 1,900 x 1,900 pixels (left), an overlapping region of 300 x 300 pixels are automatically identified in the mitochondrial channel to compute morphological features. The image is first split in a regular grid and regions with mito activity higher than a threshold is kept for quantification. **(C)** Principal component analysis of the morphological features of mitochondrial regions in the 13 images of cells. Each point represents a 300 x 300-pixel region in the mito channel. Points in blue are mito regions from wild-type fibroblasts, while points in red are mito regions from heteroplasmic fibroblasts. The principal components 4, 5, 6 and 9 were found to be significantly different between the two populations of cells and are shown in this figure. **(D)** Distribution of correlations between regions and single cells in three groups: wild-type self-correlation (WT-WT), heteroplasmic self-correlation (HP-HP), and wild-types heteroplasmic cross-correlation (WT-HP). A) Using region-based correlations. The green distribution (HP-HP) presents shifts in both tails with respect to the orange distribution (WT-HP), indicating a difference between heteroplasmic mito regions and wild-type regions. The blue distribution (WT-WT) has a significant peak at zero and at the right tail, highlighting the differences with respect to cross-correlations (WT-HP). The region-level correlations were found to have statistically significant differences. B) Distribution of similarities at the single-cell level. The blue (WT-WT) and green (HP-HP) distributions have tails to the right that indicate differences with respect to the orange (WT-HP) distribution. These differences were found to be non-significant.

In contrast, at the single cell level, the intercellular results were not statistically significant. This lack of statistical power at the single-cell level is likely due to the stochastic nature of heteroplasmy, which makes it difficult to parse out morphological differences. However, intracellular mitochondrial region analysis has the potential to enhance statistical power by highlighting subcellular morphological differences in mitochondria when individual cells have both wild type mtDNA and variant mtDNA.

### Biochemical analysis of mitochondrial enzymatic activity and mitochondrial respiration

To determine if the differences in mitochondrial morphology in heteroplasmic m.12148T>C carrying cells also correlates with mitochondrial function, we analyzed mitochondrial respiration and enzyme activity. In investigating electron transport chain (ETC) enzyme kinetics, Complexes I and IV activities were assayed because they contain a very high concentration of histidine, which could reflect altered mt tRNA^His^ function in m.12148T>C carriers. To assess Complex I and Complex IV activity, four lines were derived directly from proband fibroblasts, two with zero or near zero mutation burden and two lines with high heteroplasmic load of m.12148T>C (60-80%). These lines were clonally expanded from single cells. Complexes isolated from whole cell extracts and normalized for total protein and enzymatic activity were then assessed by ELISA. Heteroplasmic lines carrying m.12148T>C showed significantly less Complex I activity when compared to isogenic control lines (p= 0.0030; ** one Way Anova) (**Figure 4A**). This trend was also evident with Complex IV activity (**Figure 4B).** Lines with high mutational load had significantly lower Complex IV activity compared to isogenic control lines. The isogenic design of this experiment, requisite protein normalization, and the above-mentioned results confirmed that this m.12148T>C tRNA-His has a detrimental effect on ETC kinetics in isolated experiments. We then aimed to measure metabolic deficits in live culture.

**Figure 4:**
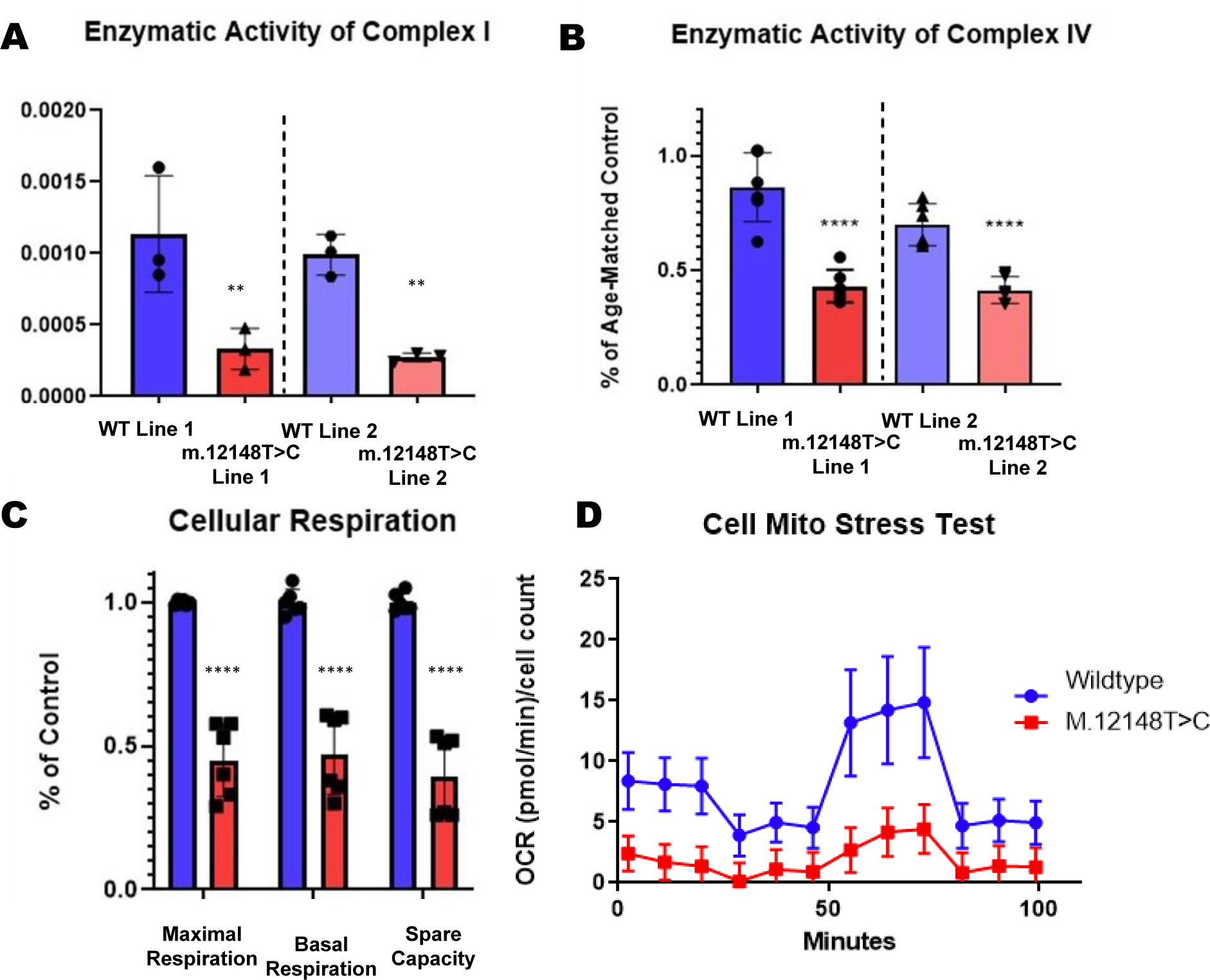
Biochemical and Mitochondrial Analysis of WT and M.12148T>C cell lines highlights deficits in Complex I& IV activity and cell respiration. **(A)** Enzymatic Activity of Complex I in Clonally derived patient fibroblast lines with determined heteroplasmy (Homo (-) 0%, and Hetero (+/-) +60%) Homoplasmic lines had higher activity than heteroplasmic lines, (p= 0.0030 ** One Way Anova). **(B)** Complex IV activity in clonally derived patient fibroblast lines with determined heteroplasmy Control (-) 0%, and Hetero (+/-) +60%) were expanded and harvested to determine Complex IV activity. Homoplasmic lines had higher activity than heteroplasmic lines, (P value <0.0001 P value summary **** One-way Anova). **(C)** Clonally derived fibroblast lines seeded for Seahorse mitochondrial Stress test, heteroplasmic lines showed lower respiration in all measured categories when compared to control Unpaired t test P value <0.0001 P value summary ****. **(D)** Single experiment representative Mitochondrial Stress test.

To investigate the metabolic implication of m.12148T>C tRNA-His mutation, we performed mitochondrial stress testing using the Agilent Seahorse platform. Assaying the oxygen consumption rates (OCR) yielded changes in respiratory capacities, with heteroplasmic lines having the lowest basal respiratory capacity: 57% of the wildtype line. The measured OCR from ATP production in heteroplasmic lines ranged from 43%-66%, relative to isogenic WT cells. Additionally, the maximum respiratory capacity in these heteroplasmic (HP) lines was 50-60% that of WT (**Figure 4C-D**). Overall, these findings suggest that the m.12148T>C mutation has an impact on mitochondrial respiration and provides further evidence for the potential pathogenic impacts for carriers.

### Heteroplasmic changes in differentiated iPSC lines

We then sought to further characterize this novel mutation in disease-relevant cells, particularly those that may reflect the disease state exhibited by our patient cohort. We aimed to understand how the heteroplasmic state of m.12148T>C changes through differentiation into various tissues, which may inform as to how this mutation distributes in disease-affected tissues and whether some tissue types are gaining a greater mutation load than others. To accomplish this, we created iPSC derived lines from all sampled tissues and isolated cells including PBMCs, fibroblasts, and cells collected in urine sediment. All tissue-isolated cells were capable of reprogramming to iPSCs (**Figure 5A**) and could maintain the m.12148T>C mutation. Furthermore, fibroblast-derived iPSCs could express a range of heteroplasmy, with some expanded iPSC colonies forming lines with no detectable/low levels of heteroplasmy and others forming lines with >60% heteroplasmy. iPSC lines derived from renal epithelium and PBMCs did not provide the range of heteroplasmy needed to create isogenic iPSC lines. As a result, we focused primarily on differentiating isogenic clones derived from fibroblasts for tissue relevant experiments.

**Figure 5:**
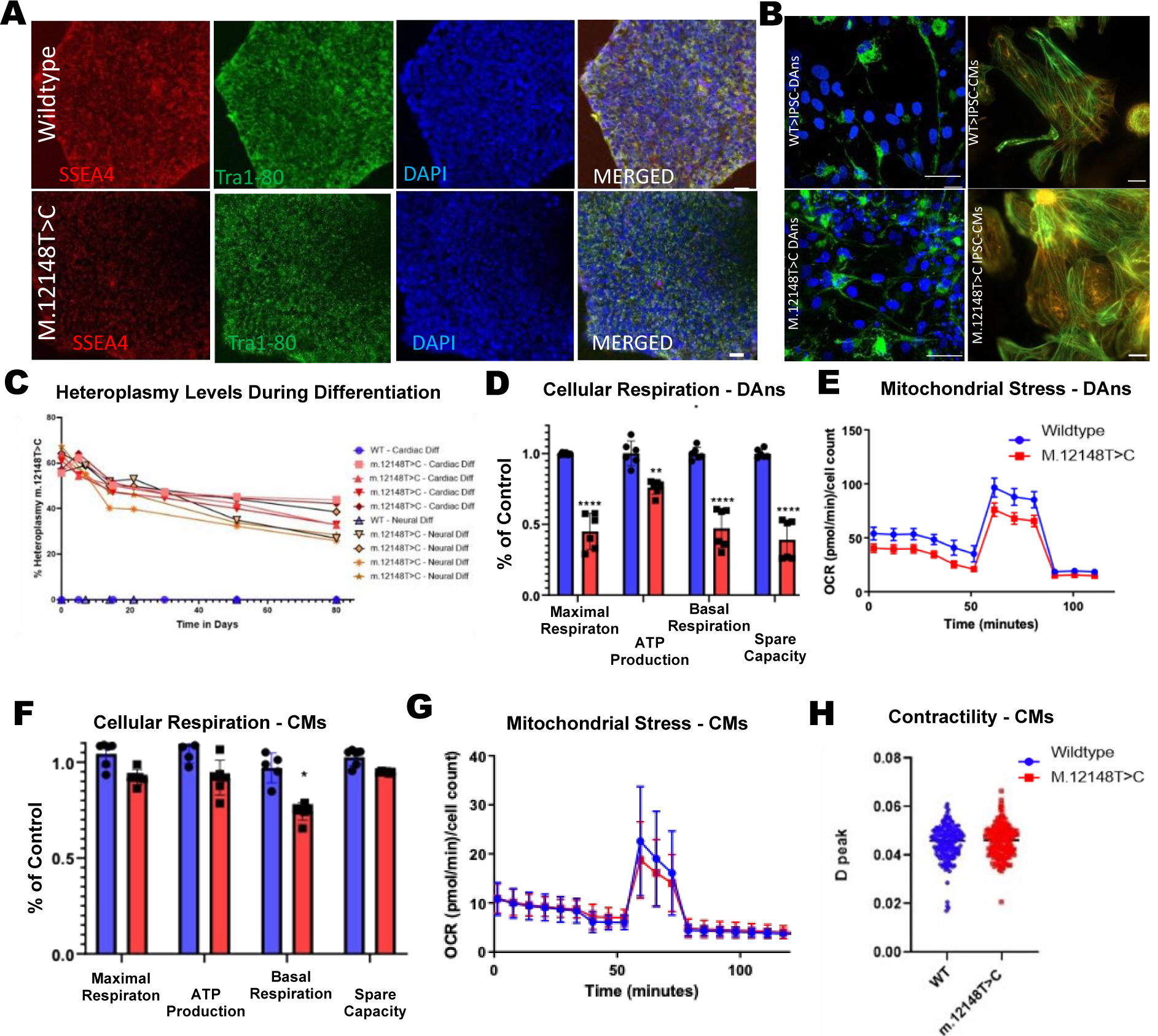
IPSC derived lines decrease in heteroplasmy upon passaging and differentiation. **(A)** IPSC lines derived from clonal fibroblast lines yielded pluripotent cells with varying heteroplasmy. **(B)** Neuronal and cardiac lineages. **(C)** Heteroplasmic IPSC lines decrease below disease threshold upon long term differentiation. **(D)** Cellular respiration in Dopaminergic neuronal (DAns) lineages shows lower respiration in heteroplasmic lines in all categories measured: Basal Respiration ATP production, Maximal Respiration and Spare Capacity. Unpaired t test P value 0.0011 P value summary** Significantly different **(E)** Single experiment representative of Mitochondrial Stress test in DAns. **(F)** Cellular respiration in patient derived Cardiomyocytes (CMs) show respiration is lower in heteroplasmic lines only in Maximal Respiration Unpaired t test P value 0.0307 P value summary * Significantly different (P < 0.05). **(G)** Single Experimental representative graph of mitochondrial stress. **(H)** Contractility of isogenic lines show no significant differences in contractility (student t test) p>.05.

### Patient derived lines recapitulate phenotype in affected and unaffected cell types

To determine the effects of differentiation to disease relevant tissue on heteroplasmy levels and cellular function, we differentiated cells towards neuronal and cardiac lineages **(Figure 5B).** We found that differentiation towards these lineages resulted in a reduction of heteroplasmy level, to around 40% heteroplasmy **(Figure 5C).** Nonetheless, to determine if these levels have a functional impact on neuronal and cardiac derived cells-particularly when compared to counterpart isogenic control derived cells-we assayed cells for oxygen consumption rates. This showed that neuronal derived lines maintained their disease relevant phenotype, as seen in the parent fibroblasts, with reduced oxygen consumption rates across all parameters in mitochondrial stress test assays (**Figure 5 D-E**). In contrast, cardiac-derived lines showed milder reductions, with decreases only evident in maximal respiration responses (**Figure 5F-G**), and no effect on contractility function **(Figure 5H).**

## DISCUSSION

The m.12148T>C is a rare/novel mutation, first reported in 2013 in a child with developmental delay, optic atrophy, hearing loss, and myopathy^10^. However, the m.12148T>C variant was not investigated beyond this clinical case report, thus has not yet been designated as a confirmed pathogenic mutation. This variant is in the D arm stem loop of the mt-tRNA for histidine presenting as a single nucleotide modification from the reference mtDNA genome and is presumed to disrupt base pairing of the stem loop structure and thus a potential alteration in aminoacyl-tRNA synthetase activity^30,31^ but had yet to be proven as a mitochondrial function disrupting event. We provide expanded data on this novel mutation and present a parent and offspring carrying the m.12148T>C mutation, with optic, aural, renal and neurologic symptoms, along with evidence of functional impacts of this mutation in relevant patient-specific cell types and in biochemical assays.

In light of compelling clinical evidence for a mitochondrial disease state, we then confirmed mtDNA mutation load through genetic sequencing, biochemical, and image analysis in different tissues. We also focused our efforts on determining the heteroplasmic status, as the affected tissue systems in our parent/sibling cohort presented with different degrees of disease severity, a potential reflection of varying mutation load. To this end, we measured the genetic prevalence and heteroplasmic state of m.12148T>C in samples from different tissues including blood, skin, and urine, by examining both bulk level and single-cell levels of the m.12148T>C mutation in different tissues.

In fact, we found heteroplasmy was distinctly different across blood, skin, and urine samples from the daughter (proband) who presented with the most severe disease state within our cohort. Specifically, we found that skin and urine samples had the highest level of heteroplasmy of m.12148T>C, which correlated with distinctly different mitochondrial morphologies in fibroblasts carrying m.12148T>C compared to age matched control cells. Furthermore, by assessing single cell heteroplasmy in these tissues we observed that different tissues have different heteroplasmy of m.12148T>C, a finding that was previously masked by bulk cell approaches. Specifically, we found that PBMCs and urine derived cells had a tight distribution of heteroplasmy, while fibroblast cells contained a large variation in heteroplasmy with cells ranging from a small number of cells with no detectable mutation to a population that was largely homoplasmic for the m.12148T>C mutation. This variation of fibroblast heteroplasmy is consistent and possibly reflective of both the degree of severity and the diversity of symptoms found in the neurological tissue in our proband, given that fibroblasts and neurons are both embryologically derived from ectoderm^24^.

Additionally, these single cell approaches were critical in assessing the causal effects of m.12148T>C on cell function, as we were able to generate isogenic controls (referred to as wild type) and isolate any potential effects of detectable nuclear variants from cells carrying the suspected disease causing mtDNA mutation. Having access to fibroblasts and isogenic control cells from the proband also provided the unique opportunity to develop iPSC lines that could be differentiated into neuronal and cardiac cell-types, tissues with similar metabolic demands as those expressing disease symptoms within our proband (neural), as well as tissues commonly associated with mitochondrial disease (cardiac). Isogenic iPSC-derived cardiac cell lines showed less of a difference in metabolic stress testing, and in contractility assays. This observation could be due to the nature of this mutation and cardiac tissues, as none of our cohort presented with cardiac related symptoms, nor has it been associated with cardiac dysfunction previously. Another possible explanation might be the influence of media factors compensating for any metabolic dysfunction. In all cases when compared to isogenic control cells, whether in fibroblasts or iPSC-derived neurons, m.12148T>C mutation carrying cells experienced diminished ETC enzyme activity and diminished respiration capacities.

Multi-tissue biochemical analysis, when taken together with genetic analysis, imaging data and modeling human tissues using patient derived iPSCs, all point towards the propensity for m.12148T>C to alter normal mitochondrial status. Furthermore, layering these findings onto the detection of this mutation only in the subjects experiencing clinically impactful and deleterious symptoms within our cohort, puts forth a strong argument to now designate m.12148T>C as a “confirmed pathogenic” mtDNA mutation rather than “possibly pathogenic” mtDNA mutation. Of course, it should be noted that this does not mean that m.12148T>C is definitively causative of a particular disease state when detected in a patient, but rather that it cannot be ruled out as a source of potential disease when found.

Ruling in a mtDNA mutation as disease causing is not trivial, and to a large degree, as demonstrated by our findings, is dependent on the heteroplasmic state of a mutation. The reason heteroplasmy disseminates differently across tissues and within individual cell types is not entirely known as there may be multiple factors at play, such as different developmental selection events^8,25,26^. These events may include different metabolic and growth pressures that may drive alternate levels of mutations within tissues, further modified by cellular heterogeneity within tissue types. For example, dermal fibroblast-derived cells have a mixture of heterogenous cell types which move between states of rapid migration/growth^27,28^, possibly lending a wide distribution of heteroplasmy states as seen in our results^7,29^. It is also possible that exposures to environmental factors *in utero* or post-natally may increase or decrease heteroplasmy in an individual, explaining drastic differences in heteroplasmy and disease manifestation across and within generations^30^. In our cohort, the parent had a low level of heteroplasmy, yet her first daughter (proband) subsequently propagated an elevated level of heteroplasmy resulting in significant symptoms. In contrast, the parent’s second pregnancy (proband’s sister) resulted in no transmission of the mutated gene. While the reasons for heteroplasmic differences across cell lineages are not fully understood, these phenomena need to be appreciated in clinical assessment of mitochondrial disease. A high mutation load in a small but relevant cell population might be masked in bulk sequencing runs of non-relevant cell populations and lead to a false negative result when mitochondrial disease is suspected.

As demonstrated here, through in-depth characterization of this novel mitochondrial transfer RNA (mt-tRNA) syndrome we highlight the importance of multiple tissue site sampling and sequencing with single cell analysis. We also highlight the utility of detecting mitochondrial DNA mutations and heteroplasmy at a single cell level and for single cell clonal expansion, as it allows for the generation of isogenic iPSCs that reflect *in vivo* disease characteristics. The combination of the data from our molecular and cellular investigations along with the clinical correlation with our patient cohort provides strong evidence for the reclassification of m.12148T>C as a mt-tRNA-his mutation from “possibly pathogenic^11^” to “pathogenic,” thus adding to the number of “confirmed pathogenic” mt-tRNA mutations and to the knowledge base towards a better understanding of mitochondrial diseases. This study also presents a set of methods and techniques that could be used for mitochondrial disease diagnosis when mitochondrial heteroplasmy is suspected, and perhaps for consideration in future mitochondrial targeted therapeutics and/or clinical trials.

## Web Resources

Phenolyzer - https://phenolyzer.wglab.org/

Burrows-Wheel Aligner - https://bio-bwa.sourceforge.net/

Picard Tools - https://bio-bwa.sourceforge.net/

Genome Analysis Toolkit - https://gatk.broadinstitute.org/hc/en-us

Deep Profiler - https://github.com/cytomining/DeepProfiler

## Data Availability

All data produced in the present study are available upon reasonable request to the authors

## Acknowledgments

This work was supported by an unrestricted grant from Research to Prevent Blindness and NEI P30EY026877, as well as a Post-Doctoral Fellowship from United Mitochondrial Disease Foundation SPO #208148.

## REFERENCES

1. Cree, L. M., Samuels, D. C. & Chinnery, P. F. The inheritance of pathogenic mitochondrial DNA mutations. Biochim. Biophys. Acta 1792, 1097–1102 (2009).

2. Schon, E. A., DiMauro, S. & Hirano, M. Human mitochondrial DNA: roles of inherited and somatic mutations. Nat. Rev. Genet. 13, 878–890 (2012).

3. McFarland, R., Elson, J. L., Taylor, R. W., Howell, N. & Turnbull, D. M. Assigning pathogenicity to mitochondrial tRNA mutations: when ‘definitely maybe’ is not good enough. Trends Genet. 20, 591–596 (2004).

4. Kujoth, G. C., Bradshaw, P. C., Haroon, S. & Prolla, T. A. The role of mitochondrial DNA mutations in mammalian aging. PLoS Genet. 3, e24 (2007).

5. DiMauro, S. & Hirano, M. Mitochondrial encephalomyopathies: an update. Neuromuscul. Disord. 15, 276–286 (2005).

6. Lott, M. T. et al. MtDNA variation and analysis using Mitomap and Mitomaster. Curr. Protoc. Bioinformatics 44, 1.23.1–26 (2013).

7. Wallace, D. C. & Chalkia, D. Mitochondrial DNA genetics and the heteroplasmy conundrum in evolution and disease. Cold Spring Harb. Perspect. Biol. 5, a021220 (2013).

8. Krjutškov, K. et al. Tissue-specific mitochondrial heteroplasmy at position 16,093 within the same individual. Curr. Genet. 60, 11–16 (2014).

9. Wang, J., Venegas, V., Li, F. & Wong, L.-J. Analysis of mitochondrial DNA point mutation heteroplasmy by ARMS quantitative PCR. Curr. Protoc. Hum. Genet. Chapter 19, Unit 19.6. (2011).

10. Tang, S. et al. Transition to next generation analysis of the whole mitochondrial genome: a summary of molecular defects. Hum. Mutat. 34, 882–893 (2013).

11. Wood, E. H., Kreymerman, A. & Randhawa, S. A new mitochondrial disease: MICHRED “‘Mitochondrial disorder with Intracranial Calcification, REnal disease, REtinopathy, and Deafness.” Invest. Ophthalmol. Vis. Sci. 59, 6052–6052 (2018).

12. Zhou, T. et al. Generation of human induced pluripotent stem cells from urine samples. Nat. Protoc. 7, 2080–2089 (2012).

13. Feyen, D. A. M. et al. Metabolic maturation media improve physiological function of human iPSC-derived cardiomyocytes. Cell Rep. 32, 107925 (2020).

14. Sharma, A. et al. High-throughput screening of tyrosine kinase inhibitor cardiotoxicity with human induced pluripotent stem cells. Sci. Transl. Med. 9, (2017).

15. Sharma, A. et al. Use of human induced pluripotent stem cell-derived cardiomyocytes to assess drug cardiotoxicity. Nat. Protoc. 13, 3018–3041 (2018).

16. Wang, K., Li, M. & Hakonarson, H. ANNOVAR: functional annotation of genetic variants from high-throughput sequencing data. Nucleic Acids Res. 38, e164 (2010).

17. Davydov, E. V. et al. Identifying a high fraction of the human genome to be under selective constraint using GERP++. PLoS Comput. Biol. 6, e1001025 (2010).

18. Rentzsch, P., Witten, D., Cooper, G. M., Shendure, J. & Kircher, M. CADD: predicting the deleteriousness of variants throughout the human genome. Nucleic Acids Res. 47, D886– D894 (2019).

19. Ioannidis, N. M. et al. REVEL: An ensemble method for predicting the pathogenicity of rare missense variants. Am. J. Hum. Genet. 99, 877–885 (2016).

20. Koch, L. Exploring human genomic diversity with gnomAD. Nature reviews. Genetics vol. 21 448 (2020).

21. Stelzer, G. et al. VarElect: the phenotype-based variation prioritizer of the GeneCards Suite. BMC Genomics 17 Suppl 2, 444 (2016).

22. Caicedo, J. C. et al. Data-analysis strategies for image-based cell profiling. Nat. Methods 14, 849–863 (2017).

24. Pawlowski, N., Caicedo, J. C., Singh, S., Carpenter, A. E. & Storkey, A. Automating morphological profiling with generic deep convolutional networks. bioRxiv (2016) doi:10.1101/085118.

24. Abdullah, A. I., Pollock, A. & Sun, T. The path from skin to brain: generation of functional neurons from fibroblasts. Mol. Neurobiol. 45, 586–595 (2012).

25. Li, M., Schröder, R., Ni, S., Madea, B. & Stoneking, M. Extensive tissue-related and allele-related mtDNA heteroplasmy suggests positive selection for somatic mutations. Proc. Natl. Acad. Sci. U. S. A. 112, 2491–2496 (2015).

26. Zhang, H. et al. Mitochondrial DNA heteroplasmy is modulated during oocyte development propagating mutation transmission. Sci. Adv. 7, eabi5657 (2021).

27. Stunova, A. & Vistejnova, L. Dermal fibroblasts-A heterogeneous population with regulatory function in wound healing. Cytokine Growth Factor Rev. 39, 137–150 (2018).

28. Sorrell, J. M., Baber, M. A. & Caplan, A. I. Human dermal fibroblast subpopulations; differential interactions with vascular endothelial cells in coculture: nonsoluble factors in the extracellular matrix influence interactions. Wound Repair Regen. 16, 300–309 (2008).

29. Floros, V. I. et al. Segregation of mitochondrial DNA heteroplasmy through a developmental genetic bottleneck in human embryos. Nat. Cell Biol. 20, 144–151 (2018).

30. Godschalk, R. W. L., Yauk, C. L., van Benthem, J., Douglas, G. R. & Marchetti, F. In utero exposure to genotoxicants leading to genetic mosaicism: An overlooked window of susceptibility in genetic toxicology testing? Environ. Mol. Mutagen. 61, 55–65 (2020).

28. Hui Yang, Peter N. Robinson, Kai Wang. Phenolyzer: phenotype-based prioritization of candidate genes for human diseases. Nature Methods, 12:841–843 (2015).

29. Sharma A, Burridge PW, McKeithan WL, Serrano R, Shukla P, Sayed N, Churko JM, Kitani T, Wu H, Holmström A, Matsa E, Zhang Y, Kumar A, Fan AC, Del Álamo JC, Wu SM, Moslehi JJ, Mercola M, Wu JC. High-throughput screening of tyrosine kinase inhibitor cardiotoxicity with human induced pluripotent stem cells. Sci Transl Med. 2017 Feb 15;9(377):eaaf2584. doi: 10.1126/scitranslmed.aaf2584. PMID: 28202772; PMCID: PMC5409837.

30. Ruth Belostotsky, Yaacov Frishberg & Nina Entelis (2012) Human mitochondrial tRNA quality control in health and disease, RNA Biology, 9:1, 33–39, DOI: 10.4161/rna.9.1.18009

31. Abbott JA, Francklyn CS, Robey-Bond SM. Transfer RNA and human disease. Front Genet. 2014 Jun 3;5:158. doi: 10.3389/fgene.2014.00158. PMID: 24917879; PMCID: PMC4042891.

